# Low Dose Regimens of BNT162b2 mRNA Vaccine Exceed SARS-Cov-2 Correlate of Protection Estimates for Symptomatic Infection, in those 19-55 Years of Age

**DOI:** 10.1101/2021.03.06.21253058

**Authors:** Graham Jurgens

## Abstract

**Background:** An exact correlate of protection (CoP) is not yet known for symptomatic COVID-19. However, it is still possible to show a new vaccine regimen exceeds an unknown CoP, provided the regimen shows an equivalent or greater immunological response in all measured indicators relative to the immunological response elicited by a clinically proven vaccine regimen. The principle of comparing immunogenicity between regimens is what the FDA, EMA, and Access Consortium use to authorize modifications to the vaccines for VOC, without requiring clinical efficacy studies before implementation. It is logical to apply the same principle to modifying vaccine doses if the data is available to do so. A two dose 30ug regimen of BNT162b2 has strong clinical evidence of efficacy, as does a single dose 30 ug regimen. The immunological markers for these regimens have been profiled in detail in Phase 1 and 2 trial data.

**Methods:** The immunological profile (including binding antibodies, viral neutralization, cytokine profiles, and CD4 and 8 expansion) of the 2 dose 30ug BNT162b2 vaccine is examined, referred to as a highly conservative CoP estimate. The single dose 30 ug BNT162b2 immunological profile is also examined, a tenable CoP estimate. Data from the phase 1 and 2 trials are examined to see if alternate regimens meet or exceed the level of each immune marker measured, relative to the regimens listed above that have proven clinical efficacy.

**Results:** For adults aged 19-55, a 2 dose 10ug BNT162b2 regimen elicits a comparable response to the standard 30 ug dose for each immune indicator, with viral neutralization nearly an order of magnitude greater than the tenable CoP estimate. Similarly, a single dose 10ug BNT 162b2 regimen or a two dose 1ug BNT 162b2 regimen equals or exceeds the immunogenicity of a single 30 ug dose.

**Conclusion:** If it is reasonable for the FDA, EMA, and Access Consortium to approve vaccine modifications without a clinical trial based on immunogenicity data, three alternate low dose regimens were identified that meet the requirements of having comparable immunogenicity relative to a protocol that has proven clinical efficacy. Immediate implementation of these lower dose regimens should be considered as they have major implications in alleviating vaccine supply, as well as improving vaccine side effect profile, and lowering total cost of vaccination.

## Background

Vaccine production has been a bottleneck in administering vaccines to increase population immunity to SARS-Cov-2. Variants of concern (VOC) have also emerged causing both the vaccine manufacturers and public health organizations to try to prepare for these variants. The U.S. Food and Drug Administration (FDA), European Medicine Agency (EMA), and ACCESS Consortium have updated their criteria and considerations for authorizing modified vaccines ^123^(1, 2, 3). None require further large scale clinical trials as the time taken for such a study would cause a lengthy delay and undermine the ability to respond to the VOCs. Instead, all focus on comparable immunogenicity data, especially on ensuring neutralizing antibody (nAb) and binding antibodies but also acknowledging that cell mediated immunity may play a role. Authorization is conditional on the fact that the modified vaccines are based on vaccines on which clinical disease endpoint efficacy study has been conducted. This authorization process without clinical trials is similar to the process used to authorize annual influenza vaccines (3).

The two dose efficacy rate of stopping symptomatic COVID was 95% in trial data ^4^(4), with reports from Israeli health agencies Maccabi and Cialit reporting similar figures in Israel’s population ^5^(5). One dose efficacy of BNT162b2 was 92.6% in the phase 3 clinical trials 14 days after receiving the dose, with multiple sources of data globally confirming a 57-90% efficacy against symptomatic disease with higher efficacies against severe disease ^6789^(6, 7, 8, 9). Early data showed lower viral titres in those that do get infected after one dose of vaccine, which would indicate likely decreased transmission ^10^(10), and and asymptomatic infections also appear to be reduced by 4 fold ^11^(11). Natural immunity post COVID-19 also appears to be protective against subsequent disease at a 94% efficacy rate ^12^(12).

Correlates of protection (CoP) for viral immunity can differ based on the virus and can include binding antibody concentration, viral neutralization titres, a favorable cytokine profile, a certain level of CD4 or CD8 expansion, immunological memory, or a combination of the above. The threshold level for protection against asymptomatic infection may be higher that of symptomatic disease, while the threshold level against severe outcomes may be lower. An exact correlate of protection (CoP) level is not yet known for any of these outcomes for SARS-Cov-2. The majority of CoPs for previous diseases protected by vaccines are assays assessing antibody titres ^13^(13), so understandably antibody responses have been evaluated carefully for each vaccine trial. However, raw antibody titres may not be the best correlate of protection, given that high levels of antibody titres are correlated with severe Covid presentation ^14^(14). The type of antibody appears to matter rather than just the overall amount in response to Covid-19: afucosylated IgG1 antibodies in particular are associated with severe disease along with Th2 polarization of immune response ^15^(15). Low antibody titres are correlated with faster viral clearance (14), perhaps indicating the importance of cell mediated immunity and Th1 polarization of the immune system. The cytokines elicited by the mRNA vaccines are most often associated with a polarized Th1 response: IFN gamma, IL2, and TNF ^16^(16).

If a specific immune indicator defines a CoP, a new vaccine regimen being evaluated would need to demonstrate that it exceeds that specific indicator to be effective (antibody titres being very common). For an undefined CoP, it is still possible to demonstrate a new vaccine regimen exceeds the CoP, but it would be required to demonstrate an equivalent or greater immunological response in all the immune indicators considered as candidates in defining indicator of the unknown CoP, relative to the immunological profile elicited by a clinically proven vaccine regimen. This is the underlying reason why immunogenicity data is sometimes used to estimate a CoP when it is not feasible to do the clinical efficacy studies.

In the context of vaccine shortage, fractional low dosing is a strategy used to help mitigate supply limitations and has been successfully used with a number of vaccines, polio and yellow fever being two examples. ^17^(17). Fractional low dosing of vaccines often elicits an immune response equivalent to a single standard dose, and are most often studied in non elderly, non-malnourished, healthy subjects. How low of a fractional dose can be given depends on the vaccine; in the case of yellow fever, doses 50x lower than standard still led to strong immune protection where doses 100x lower did not. Despite a lower total dose of vaccine given, the duration of immunity with fractional doses can be comparable to the standard dose. ^18^(18)

## Methods

The immunological profile of two vaccine regimens that have proven clinical efficacy data were evaluated. First, the 2 dose 30ug BNT162b2 profile was examined, including antibodies, cytokine profiles, and CD4 and 8 expansion. This will be termed a highly conservative CoP estimate, because the responses seen are nearly an order of magnitude higher than the single dose profile, which also has shown strong clinical efficacy. The data was taken from the Phase 1/2 data published in the NEJM (16), a trial based in the USA, as well as an extended dataset looking at a continuation of antibody response and evaluating T-cell response, based out of Germany ^19^(19). The single dose 30 ug BNT162b2 immunological profile was also considered: This will be termed a tenable CoP estimate. Dosing regimens from the phase 1 and 2 data are examined to see if alternate regimens are equivalent to the above regimens for each immune marker measured. Only data from the 19-55 year age group was considered.

## Results

### Antibody levels and viral neutralization

Figure 1A shows the extended trial data from Pfizer/BioNTech. The authors noted that although the S1-antibodies showed a gradual decline (typical post infection or vaccination after initial proliferation and contraction), the viral neutralization titres stabilized at day 43, likely indicating selection and affinity maturations for the antibodies (19). The 30ug VNT50 is comparable to the 10ug titre from days 43 to 85. Both of the 30 ug VNT50s appear to be larger than 10 ug at day 29, but this is potentially sampling bias, as a similar two fold difference can be seen between the Germany and USA trials who received 20 ug dosing, see Fig. 1C.

**Figure 1.**
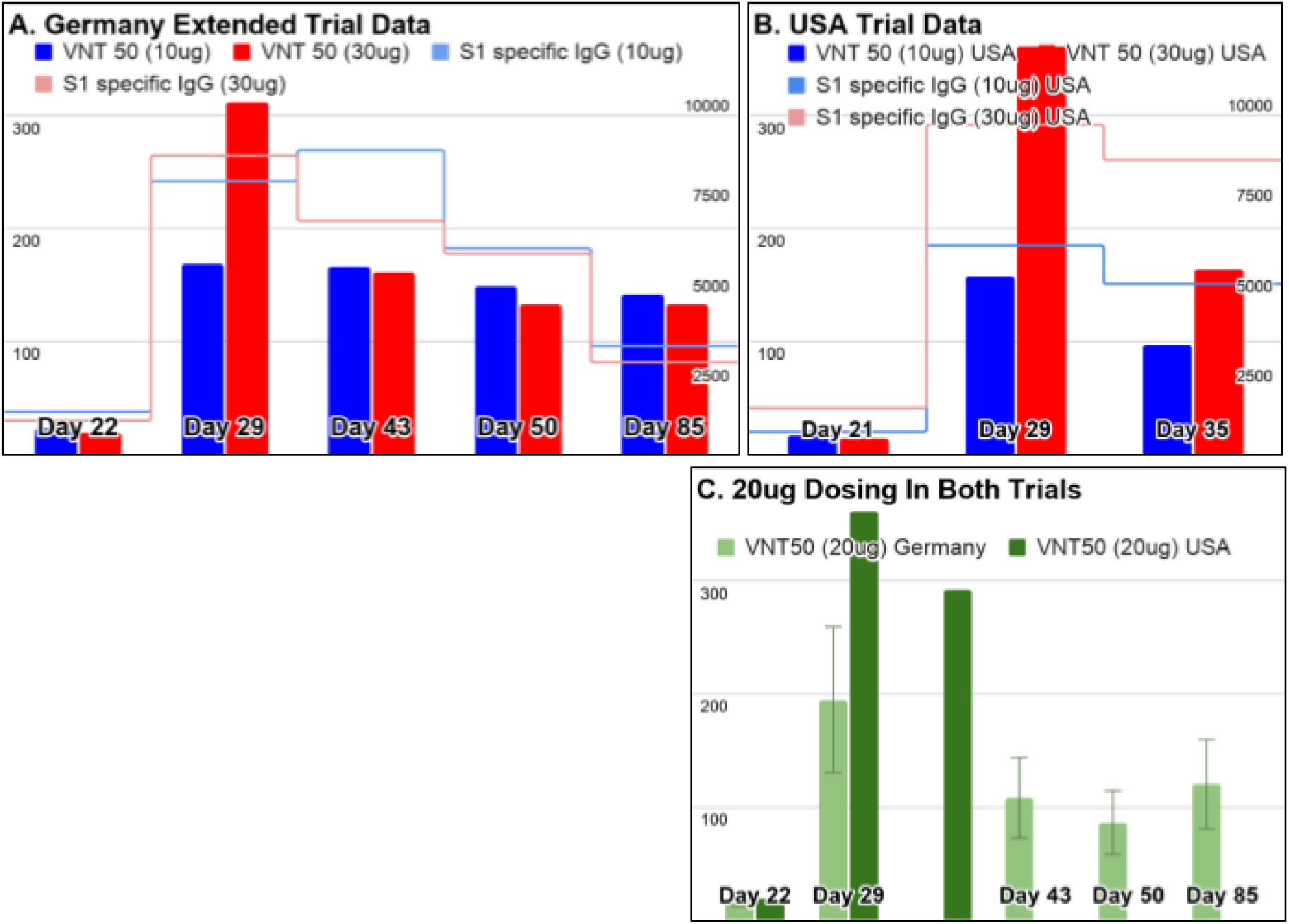
10 ug and 30ug Dosing Similar Once Adjusted for Sample Bias. Day 29 shows an increased VNT50 for 30 ug. The 10ug and 30ug VNT50s stabilize at a similar level thereafter. S1-specific andibodes are comparable throughtout, see Fig 1A and 1B. Figure C shows that even with the same given dose, a two fold difference can be seen between titres, perhaps due to sampling bias. Note the relatively low VNT50 titres before dose 2 was given at day 22, which produced a clinical efficacy against symptomatic COVID-19 of 92.6%. Left axis: neutralization titres. Right axis: antibody concentration

The S1-specific antibodies are comparable between dosing groups, with Sahin et al commenting there was “evidence of a dose level-dependent response only between the 1ug and 10 ug dose levels” and thus not between 10ug and 30ug groups (19). They also comment that the S1-antibodies strongly correlate with the VNT50. The 30ug and 10 ugs VNT50s from day 43 to 85 after stabilization appear equivalent. Additionally, the VNT50 levels after 10 and 30 ug doses are nearly an order of magnitude higher than the VNT50 for a single dose level seen on day 22 for which trial data showed clinical efficacy of 92.6% (4), which would indicate that even if there could be found a statistically significant difference in immune markers between at day 29, there is likely to be no clinical significance to that difference.

### Cytokine profile and Cellular Expansion

T-cell responses are important to consider as a CoP. The authors comment on the potential importance of cellular expansion saying “T cells could be relevant in disease control even in the absence of neutralising antibodies,” as it has been seen that mild and asymptomatic infection by SARS-Cov-2 have been cleared without seroconversion (19).

There was lots of individual variability when looking at cellular expansion. Sahin et al. comment that “Seven days after the boost with BNT162b2 at any of the doses, robustly expanded SARS-CoV-2 S-specific CD4+ T cells were detectable in all 37 participants…for dose levels ≥10 μg the magnitude of CD4+ T cell responses did not appear to be dose-dependent” and “the magnitude of the T cell responses varied inter-individually and showed no clear dose dependency. Even with the lowest dose of 1 μg BNT162b2, most of the vaccinated participants demonstrated robust expansion of CD4+and CD8+ T cells.” (19)

Despite having lower antibody concentration and viral neutralization titres compared to 30 ug dosing, 1ug dosing had robust expansion of T-cells. The robustness of T-cell expansion to low doses was also seen with the closely related BNT162b1, for which the author’s comment that they “observed no clear dose dependency of the T cell response strength within the tested dose range (1–50 μg). Even with a dose as low as 1 μg, mRNA-encoded immunogen stimulation and robust expansion of T cells was accomplished in most subjects” ^20^(20).

In summary, the expansion between 10ug and 30ug appears to be equivalent. Cell expansion with 1ug, if not quite as prominent, is “robust”. It’s important to note that T-cell responses were not assessed after just one dose of the vaccine for Pfizer. For Moderna, that data was obtained, and after one dose there is a slight increase of expansion with the majority of it happening after the second dose ^21^(21). That is likely the case here as well, and it would be reasonable to expect that one dose of 30 ug BNT162b2 would have less T-cell expansion than 2 doses of 1ug, following the trend from Moderna’s data. The T-cell responses did correlate positively with S1-binding IgG for both CD4+ and CD8+ T-cells, and the concentration of S1 IgG is higher after two 1ug doses than after a single 30ug dose, which might be mirrored by cellular expansion (19), see Fig 3.

**Figure 2:**
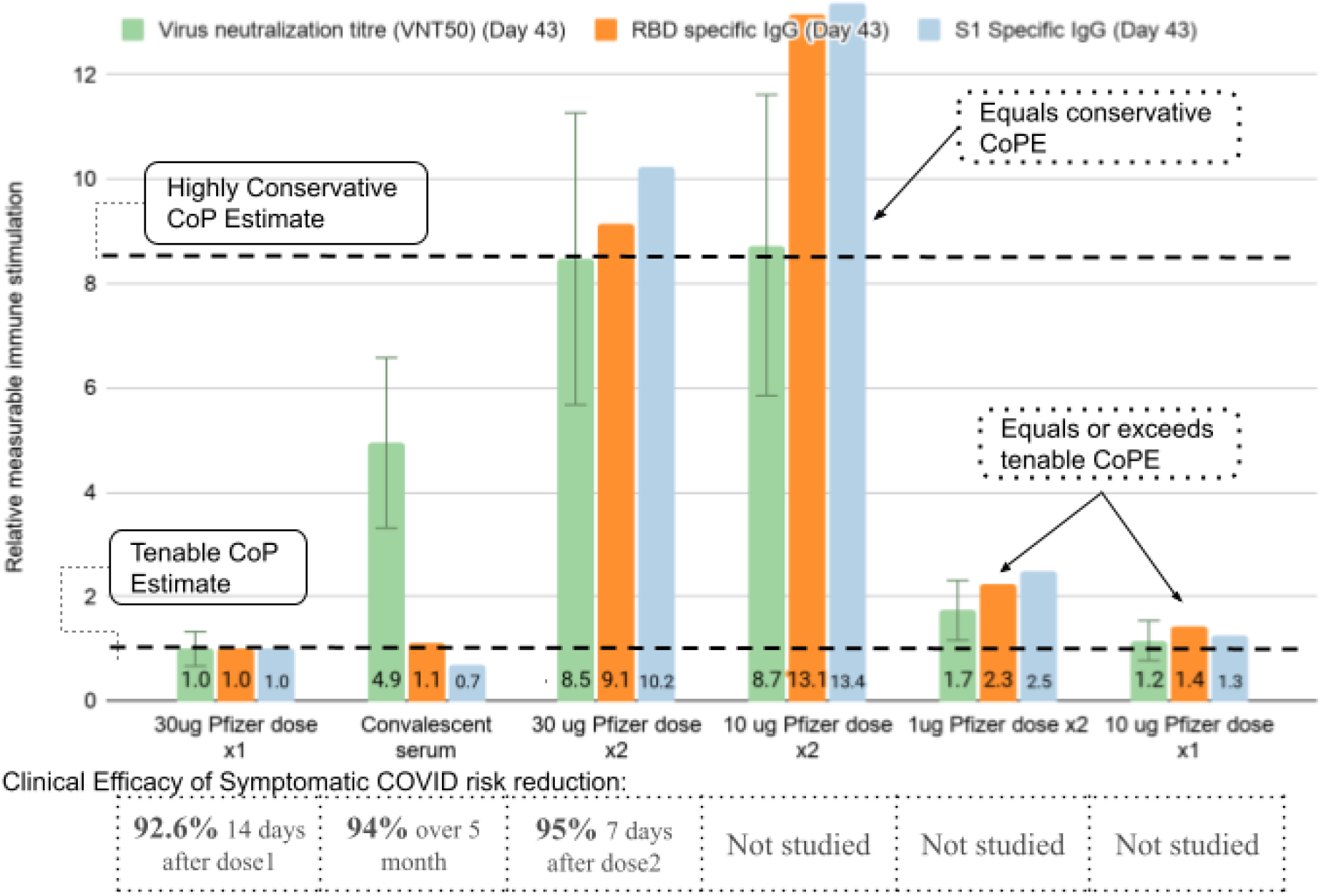
Relative BNT162b2 Antibody and Virus Neutralization Titres. Extended trial data from Germany. Neutralization titres are graphed at day 43 to allow for stabilization of titres. A single dose of BNT162b2 is set as the baseline as a tenable CoP estimate, with the other data graphed relative to that baseline. The highly conservative CoP estimate is nearly an order of magnitude higher than the tenable CoP estimate, while convalescent serum levels are in between. Two doses of 10ug is comparable to the highly conservative CoP estimate, far exceeding the tenable CoP estimate. A single dose of Pfizer at 10 ug is equivalent to the tenable CoP estimate and a two dose, 1ug Pfizer regimen exceeds the tenable CoP estimate.

**Figure 3:**
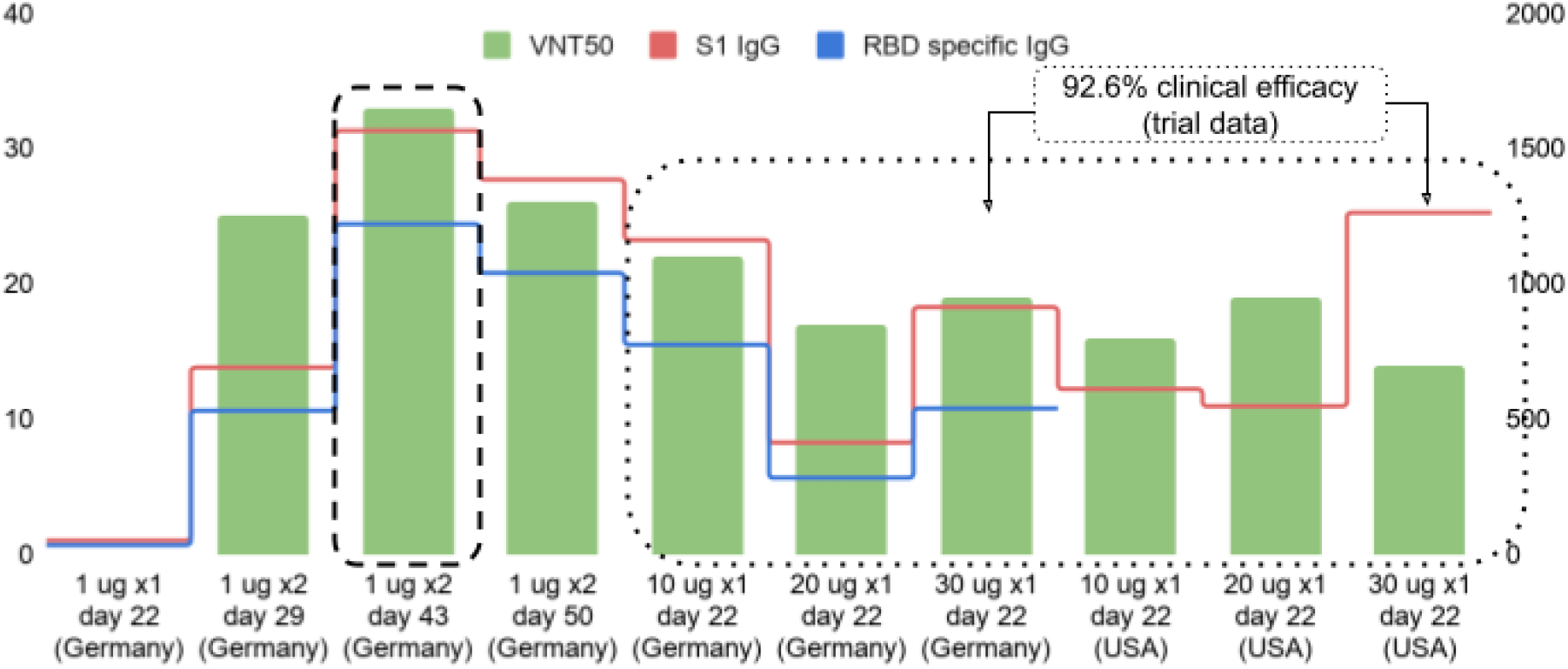
1 ug Immune Response Compared to Single Dose 10-30ug Responses. No immune response was seen after a single 1ug BNT162b2 dose. Three weeks after a second 1ug dose (dashed area), immune responses were higher than single dose regimens at a dose of 10 to 30 ug at 3 weeks (dotted area). T-cell expansion correlated with S1-lgG concentration. T-cell expansion for two dose 1ug regimen was characterized as ‘robust’.

The cytokine profile for all dosing regimens tested were similar. Expression of IFNg and IL2 were seen, with barely detectable expression of IL-4: indicating “a TH1 profile and the absence of a potentially deleterious TH2 immune response” (19).

### FDA, EMA, and ACCESS Consortium Requirements for Modified Vaccines

The FDA and EMA requirements for authorization of vaccine modifications against VOCs were likely not intended to be used to analyze altered dosing regimens efficacy against the original virus. Nevertheless, the concepts behind authorization are instructive.

The FDA states that the outlined guidelines were applicable when for the modified vaccine when “it is not feasible to conduct clinical disease endpoint efficacy studies rapidly enough to respond to the emergence of SARS-CoV-2 variants that may escape immunity conferred by prototype vaccines” (1). The modification discussed in this paper is a modification to dose rather than to structural change. Certainly the original SARS-Cov-2 is a virus of concern, and the titres elicited by the current vaccine does appear to have good efficacy against many of the VOC as well, including B117. It is not feasible to repeat another Phase III trial with the lower dose regimens discussed rapidly enough to respond to the already widespread original virus and growing B117 VOC.

The alternate vaccine sparing regimens meet all the immunogenic criteria as set out by the FDA, EMA, and ACCESS Consortium, see Table 1.

**Table 1.** Requirements For Authorizing Modified Vaccine Regimens

| FDA/EMA/ACCESS Consortium Requirement | Requirement met by 10ug dosing x 2 doses (yes/no) | Requirement met by 1 ug dosing x 2 doses (yes/no) |
| --- | --- | --- |
| Not feasible to do a large phase 3 trial rapidly enough to respond to a virus? | Yes | Yes |
| Parent vaccine already approved? | Yes | Yes |
| Vaccine made with the same process that led to clinical disease endpoint efficacy study that meets FDA success criteria? (50% reduction of disease) | Yes | Yes |
| Vaccines made by the same manufacturer and process? | Yes | Yes |
| Neutralizing antibody response rates similar (FDA: within 1.5 fold) to proven vaccine regimen to<br>A: 2 dose regimen of 30 ug?<br>B: 1 dose regimen of 30 ug? | A: Yes- equivalent once stabilized, see Figure 1.<br>B: Yes- far exceeds | A: No- reduced titres<br>B: Yes- exceeds. |
| Binding antibodies geometric mean concentration (GMC) similar (FDA: within 1.5 fold) to previous proven vaccine regimen<br>A: 2 dose regimen of 30 ug?<br>B: 1 dose regimen of 30 ug? | A: Yes, equivalent<br>B: Yes, far exceeds | A: No-reduced concentration<br>B: Yes-exceeds |
| Vaccine tested against VOC? | Yes (B1.1.7) and original strain | Yes (B1.1.7) and original strain |
| Using the dose and dosing regimen as authorized for the prototype vaccine? | No | No |
| Safety assessments completed? (may need to monitor with large sample size if concerns arise with a booster) | Yes: less reactogenic compared to standard dosing | Yes: much less reactogenic than standard dosing |

### Reactogenicity and Population Immunity Considerations

For those aged 19-55, two 10 ug doses appear to equal a highly conservative CoP estimate, and two 1ug doses appear to exceed the threshold of a tenable CoP estimate. How that might affect an individual receiving the vaccine and a population vaccinating to protect against COVID-19 also need to be considered.

On an individual level, it is clear from the safety profile of the data that 10 ug dosing has a more favorable reactogenicity profile, as illustrated in Fig. 4. In the context of emerging and spreading VOCs, manufacturers are evaluating boosting with a modified strain or using a 3rd dose of the original vaccine to help boost immunity. As there is a significant increase in reactogenicity with the second dose (especially in systemic side effects), it would be reasonable to expect the reactogenicity of a 3rd dose to be equal or higher than that of the second dose. The observed decrease in vaccine side effects from using 10 ug doses instead of 30 ug doses would likely also apply to a 3rd dose, if a 3rd dose will be required. This is important consideration not only for the individual in the context of missed days of work after receiving a booster, but also for public health as a unfavorable side effect profile may increase vaccine hesitancy ^22^(22).

**Figure 4:**
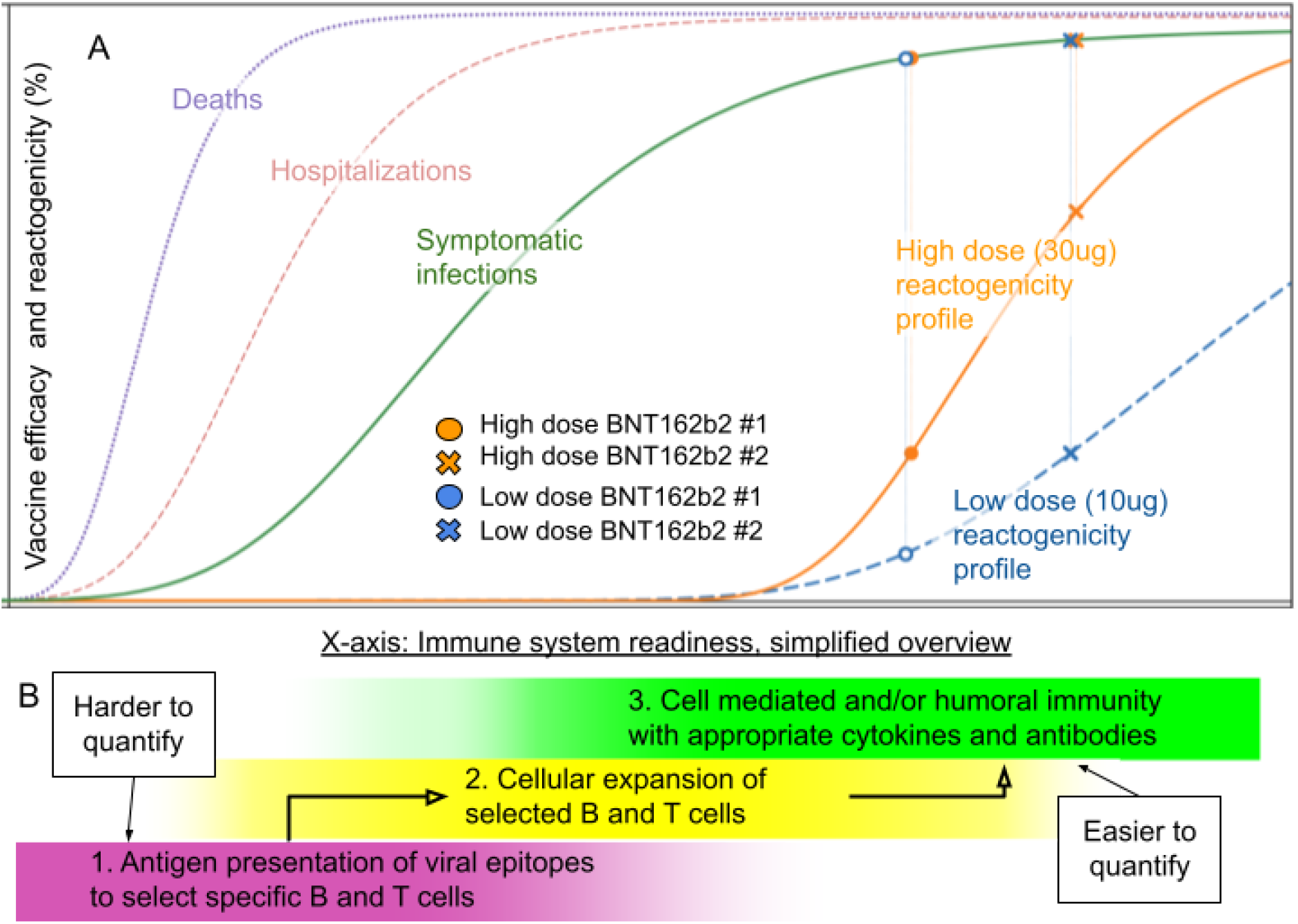
Reactogenicity with High and Low Dose Vaccine Regimens. A two dose 10ug regimen gives equivalent immunogenicity markers compared to the two dose 30 ug regimen with an improved side effect profile. All curves are estimates. Vaccine efficacy against death and hospitalizations appears to be greater than efficacy against symptomatic disease. 4B illustrates a simplified overview of the steps that the immune system takes post vaccination to prevent disease. Note that the two dose 1ug regimen of BNT162b2 is not plotted on the graph, but they have an even more favorable reactogenicity curve than 10ug, while inducing robust cellular expansion with antibody titres just above a single 30 ug dose.

More important than reducing vaccine side effects is protecting the population from disease. On a population level, using 10ug doses instead of 30ug doses for those aged 19-55 effectively triples the vaccine supply for a large portion of the population. In the context of vaccine shortage, the effect is enormous, see Fig 5.

**Figure 5:**
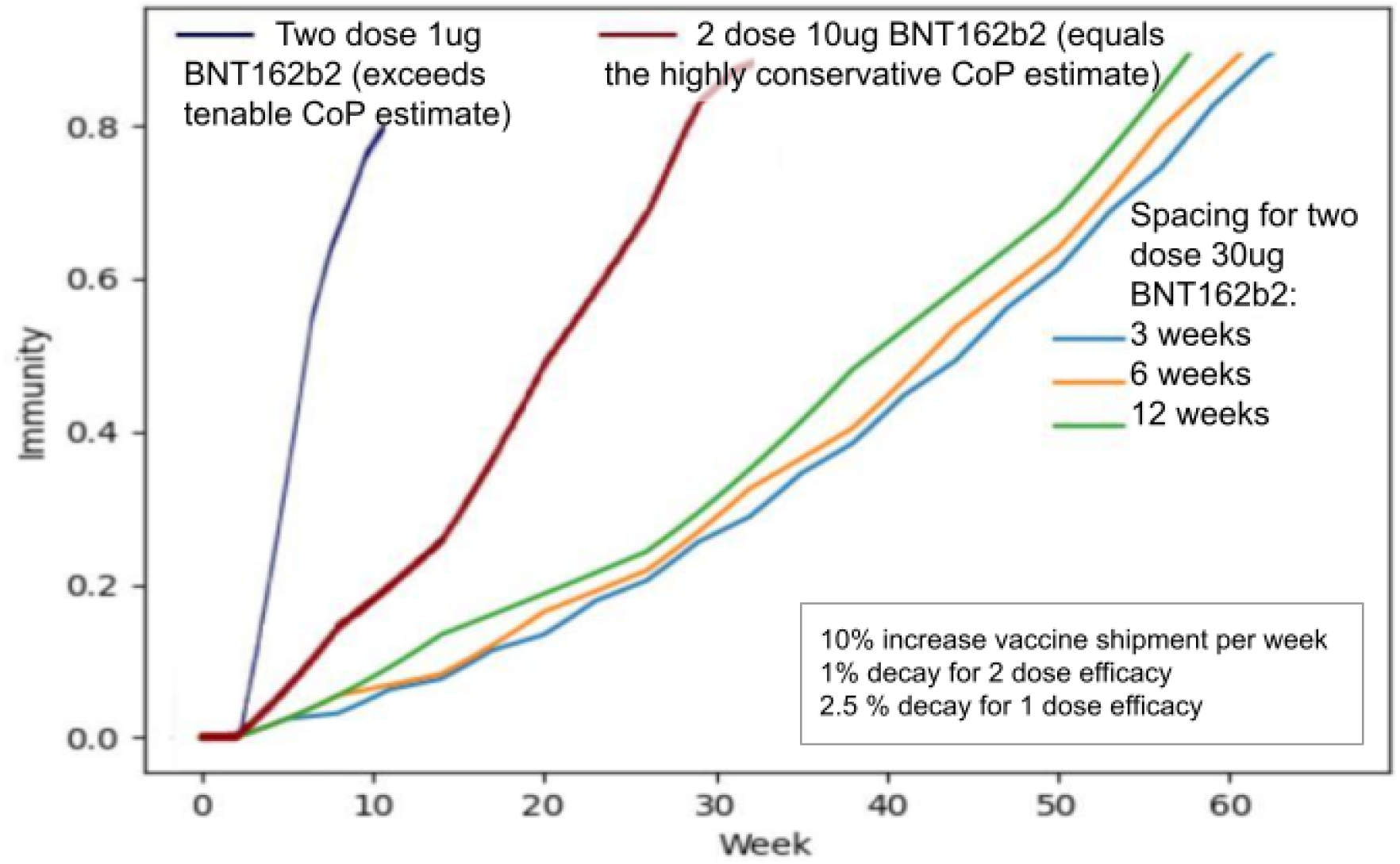
Population Immunity Modelled with Low and Ultra Low Dose Regimens. Population immunity is increased significantly from the standard two dose 30ug regimen given 3 weeks apart (blue) by delaying the second dose (orange and green), and is increased drastically by using a two dose 10ug regimen (red) and two dose 1ug regimen (navy). The two dose 10ug curve assumes 90% efficacy after to doses, while the two dose 1ug curve assumes 80% efficacy after two doses. The graph assumes no administration limit.

Figure 5 shows that tripling the number of vaccine doses by using 10ug doses (instead of 30 ug doses) for adults aged 19-55 would half the time needed to reach maximal population immunity. Using a two dose 1ug vaccine regimen would speed that process further. For countries with robust vaccine supply, the administration of vaccine would become the bottleneck rather than vaccine supply which would limit the gains modelled in Fig. 5. For countries where scarce vaccine supply is the limit rather than administration, the gains would be highly significant.

## Conclusion

Fractional dosing of Pfizer/BioNTech BNT162b2 (at 10ug per dose) appear to have near identical immune stimulation to the higher 30ug standard doses in 19-55 year olds, as measured by antibodies levels, viral neutralization titres, cytokine release profiles, and CD4 and CD8 expansion. Which one (or multiple) of these markers is the most predictive of protection and will eventually define the CoP for COVID-19 is unknown, but is not needed to be known since the 10ug dose appears to be comparable to 30 ug in each of the indicators. It follows then, that the lower 10ug dose would seem to reach a highly conservative CoP estimate and pass all of the FDA, EMA, and ACCESS Consortium immunogenicity requirements for approval for a modified vaccine. Implementation of this dosing regimen should be considered without further clinical efficacy studies. More antigen being presented to the immune system does not always equate to higher immunogenicity markers, which appears to be the case between 10 and 30 ug dosing. This is also seen in COVID-19 recovered patients who increase their VNT50 and antibody titres after one dose of vaccination, but not after the second ^23^(23). It is also seen in AstraZeneca’s trial data where a low dosing regimen led to equivalent if not greater efficacy than standard dosing ^24^(24).

It may be possible that something beyond what was clinically examined in the trial data ends up being the CoP, an as of yet unknown mechanism of protection, but it would be unexpected. The fact that the FDA, EMA, and ACCESS consortium are evaluating immunogenicity primarily based on viral neutralization and antibody titres as surrogates for efficacy when considering approving vaccine modifications would indicate that there is no clear reason to expect an unknown mechanism of protection beyond what has been studied. Neutralizing antibodies and T cell responses appear to play a role in protection. Non-human primate (NHP) studies show vaccine elicited nAb titres correlate with protection, and that the correlation was stronger when combined with diverse Fc functional antibody response ^2526^(25, 26). Other studies have shown that mild COVID-19 recovered individuals have low nAbs, but robust T-cell response with memory T-cells that could protect against future severe COVID ^27^(27).

If nAbs are important in preventing COVID-19, they may not have to be initially present at detectable levels for protection against disease: The immune system is dynamic, and as long as it has been stimulated and has SARS-Cov-2 specific memory cells, the quick reactivation and production of antibodies from memory cells may be enough to protect against disease. This would be similar to protection against Hepatitis B infection: antibodies are protective, but despite antibodies waning to undetectable levels, efficacy from the vaccine remains high ^28^(28). This is not always the case of course, an example being preventing Hemophilus influenza B (Hib) meningitis: when initial antibody levels are too low, Hib can still cause disease despite the quick production of antibodies from memory cells ^29^(29). The single dose regimen of BNT162-b2, with relatively low neutralizing antibodies yet strong efficacy, may indicate that if nAb are crucial to prevent disease, then memory cells are able to increase the nAb level quickly enough to prevent disease.

There is historical precedence using an estimated correlates of protection from NHP studies for human use, rabies vaccine development being one example ^30^(30). Lower doses in children are expected to be studied, including half and quarter dose regimens ^31^(31) Comments from one of the principal investigators in J and J and Moderna clinical trials ^32^(32) as well as from Pfizer’s investigator ^33^(33) would suggest that clinical vaccine efficacy in children will be inferred from an estimated CoP rather than observed clinical efficacy data. Yearly annual influenza vaccines are approved without clinical efficacy trials based on a CoP ^34^(34).

There are multiple benefits to using lower dosing in the 19-55 aged population. It greatly reduces vaccine supply constraint, and as long as administration of the vaccines allows, it would vastly reduce the time needed to reach a set immunity level, which would prevent disease as well as reduce the need for lockdowns and other non pharmaceutical measures.

The reactogenicity profiles are milder for the lower dose regimens, which could reduce vaccine hesitancy (22). Anecdotally there have been many reports of significant symptoms after the second dose of vaccine leading to time lost at work. Using lower doses could significantly minimize these events. This is especially important when considering the potential need for a 3rd dose of a vaccine as a booster against VOCs: the reactogenicity for dose 2 is greater than that of dose 1, and is unlikely to decrease with a 3rd dose. This could be a factor in why Moderna is only trialling 3rd doses as 50ug or less, rather than using a standard 100ug dose ^35^(35).

Beside the cost of lost time at work due to vaccine side effects, the cost of the vaccine itself is an additional benefit to lower dose vaccination regimens. Using vaccine price of 19.50 USD per vaccine dose for Pfizer ^36^(36), in using the BNT162b2 at 10ug/dose, the cost saving per person is 26 USD ($19.50×2doses/3). Using an estimate of 190 million people in the USA between ages of 19-55, and if 75% are vaccinated with a lower dose mRNA vaccine regimen at an average saving of $26 per person, a cost saving of 3.7 billion USD could be saved compared to the standard vaccination series. If a 3rd dose is required as a booster, the financial savings would further increase.

The above benefits consider implementing a two dose 10ug BNT162b2 regimen, which equals the immunological profile of an highly conservate CoP estimate. There is little reason to use the standard 2 dose 30ug regimen’s immunological profile as a CoP estimate now that there is clear clinical efficacy evidence showing that the lower immunological profile of single 30ug dose (with neutralization titres nearly an order of magnitude less) still provide strong clinical efficacy. This single dose immunogenicity profile is a tenable CoP estimate. It also is likely not the minimum threshold required for protection, but it gives at least a closer estimate of the levels that will eventually define the CoP compared to the highly conservative CoP estimate.

Multiple alternate regimens in the Phase 1/2 trials with a lower total vaccine quantity exceeded the tenable CoP estimate. Of note, the immunologic profile of 1ug doses given 21 days apart is favorable relative to one 30ug dose, showing higher levels of antibody titres and virus neutralization, with a robust expansion of CD4 and CD8 cells while showing a favorable cytokine profile. A single 10ug dose also appears to be equal to the tenable CoP.

The benefits to using a two dose 1ug regimen are magnified even more so than using a two 10ug dose regimen as described above. The side effect profile is even more favorable at 1ug doses. It would reduce vaccine cost per dose from 19.50 USD to 0.65 USD, making it much more economically feasible for less affluent countries. It would greatly reduce vaccine supply as a bottleneck for vaccination in most countries. The time it takes for less affluent countries to access to the vaccine would be shortened further.

One concern about the vaccine efficacy of a single dose 30ug regimen is the unknown duration of that immunity, and that concern would also apply to the two dose 1ug vaccine regimen. There has been no indication from countries delaying second doses (like the UK and Canada) that one dose efficacy has significantly waned, and the two dose 1ug vaccine would be likely to follow a similar trajectory. If immunity were to wane after a two dose 1ug regimen then a third dose would be required to optimize protection. Of note, the potential need for a third dose may also be true for the standard regimen. If T-cell response is a surrogate for determining the duration of immunity, then the duration of immunity between 30ug and 1ug dosing regimens might be similar, given they both elicit robust cellular responses, but if antibody or neutralization is a surrogate for duration of protection, then the standard dose with its order of magnitude higher antibody and neutralization response would indicate longer duration of immunity.

Using a single 10ug dose instead of a 2 dose 1ug regimen would have the advantage of requiring one less inoculation, but the disadvantage to this regimen is that it uses 5 times the amount of vaccine as the two dose 1ug regimen. As noted in Figure 3, there was no measurable immune response after a single 1 ug inoculation, so a two dose 1 ug regimen would be unlikely to have significant protection until 7 days after the second dose. A single 10 ug dose regimen, in contrast, would have some protection as early as 14 days after the first dose, two weeks earlier than the two dose 1 ug regimen.

Another important consideration is vaccine supply and the capacity to administer the vaccine. For a region with a robust vaccine supply that is less cost conscious, the capacity to administer the vaccine might be a bottleneck rather than total vaccine supply. If that is the case, using a single dose of 10ug, followed by a second 10ug dose if needed, is logical. For countries that are more cost conscious or where supply is the major limitation to vaccination, then stretching each dose of vaccine as far as possible with a 2 dose 1ug regimen might be favored. The equity of vaccine distribution between wealthy and poor countries appears mostly to be an afterthought in practise ^37^(37), but it is important to note that the less vaccine that is required for a vaccine rich country to protect its population should mean that vaccine poor countries will have access to vaccines sooner. Only once global immunity has increased will viral replication decrease and the risk of new variants will decrease along with it.

A similar evaluation of Moderna mRNA-1273 data would show that 50ug per dose appears equivalent to 100ug dosing ^38^(38) and by similar reasoning would also equal a highly conservative CoP estimate. As a single dose of Moderna also showed robust efficacy in the trial data, a two dose 25ug regimen exceeds a tenable CoP estimate (and it is likely that even much lower doses than 25ug would meet or exceed a tenable CoP estimate).

In summary, when a CoP is not well defined, it appears using a CoP estimate appears to be a reasonable approach, based on years of experience doing the same with other vaccines, like the yearly influenza vaccine (34). It seems generally accepted that it is reasonable to evaluate the efficacy of modified vaccines tailored to VOCs in a similar manner, and this will likely be how the SARS-Cov-2 vaccine efficacy will be evaluated in children. All of these will be implemented without clinical efficacy trials, although the efficacy of modified vaccine regimens post implementation may be monitored. In the midst of an ongoing pandemic, evaluating lower dose regimens in the same manner is logical.

The immunological profile of a two dose 10 ug of BNT162b2 regimen appears equivalent to that of the two dose 30ug regimen, which is itself a highly conservative CoP estimate. Without clinical efficacy trials, a 10ug dosing should replace the standard 30 ug dose for all adults aged 19-55, even in the context of robust vaccine supply: while the immunogenicity appears equivalent, one would gain the benefits of lower vaccine cost and a favorable reactogenicity profile for the regimen, including a 3rd dose specific to VOC if needed.

By the same rationale, in the context of vaccine shortage, a two dose 1ug regimen or a single 10 ug dose should be considered for adults aged 19-55 without clinical efficacy trials, as the immunological profiles of those regimens equal or exceed a tenable CoP estimate, further decrease costs and side effects, and most importantly reduce supply constraints. This would appear to dramatically increase the ability to augment total population immunity.

The FDA, EMA, and ACCESS consortium are not reckless authorizing vaccine modifications without clinical efficacy, provided there is comparable immunogenic data. Considering the context, it is prudent. Failure to apply the same standard in evaluating low dose vaccine regimens would appear to disregard the years of experience using CoPs with other vaccines, and would mean losing the tremendous benefits gained by minimising supply constraints.

## Supporting information

Supplementary Appendix: Using a Correlate of Protection Estimate to Predict Treatment Efficacy

## Data Availability

All data generated or analysed during this study are included in this published article (and its supplementary information files)

