## Supplementary Appendix: Using a Correlate of Protection Estimate to Predict Treatment Efficacy for "Low Dose Regimens of BNT162b2 mRNA Vaccine Exceed SARS-Cov-2 Correlate of Protection Estimates for Symptomatic Infection, in those 19-55 Years of Age"

Author: Graham Jurgens, MD BSc Microbiology and Immunology


A correlate of protection (CoP) can be defined as an immune marker statistically correlated with vaccine efficacy. When the immune marker being evaluated is the causal reason for protection, it can be referred to as a mechanistic CoP (mCoP) and when the immune marker being evaluated is not the causal agent of protection yet still predicts efficacy, it can be referred to as a non-mechanistic CoP (nCoP). The advantage to knowing a defined CoP is that it can be used to assess the protective efficacy of a treatment being evaluated without having to directly measure clinical outcomes, leading to expeditious approval and implementation of that treatment.

When it is unknown which immune marker will eventually define a CoP, or when the threshold level needed for protection is unknown for an established immune marker that correlates with protection, I propose that a "CoP estimate" (CoPE) can be used to predict treatment efficacy.

A CoPE can be defined as the collective immunological profile of a treatment proven to be clinically efficacious. The CoPE immunological profile must include all of the potential immune markers that could be expected to eventually define the unknown CoP.

If the response to a treatment being evaluated is to meet a CoPE, it would need to meet or exceed the threshold for all of the immune markers making up the CoPE immunological profile, because the specific marker that will make up the mCoP or nCoP has not yet been established.

To reach a CoP threshold, the treatment being evaluated must result in a known level of a known immune maker that predicts efficacy, while to reach a CoPE threshold, the treatment being evaluated much equal or exceed the levels of all the immune markers that make up the CoPE immunological profile.

An example may help illustrate: Vaccine A produces strong clinical efficacy against a disease, but a CoP has not yet been defined, that is, the protective immune marker and level of that marker needed to predict protection is unknown. If the only potential immune markers expected to define the eventual mCoP or nCoP include binding antibodies, neutralization titres, CD4 and CD8 expansion, cytokine profiles, or a combination thereof, then those immune markers make up the CoPE's immunological profile. As long as Vaccine

B equals or exceeds the levels of all the above listed immune markers relative to Vaccine A, it can be surmised that Vaccine B meets the CoPE, and will be efficacious against disease, even in the absence of directly measured clinical outcomes. See the Supplementary Index for how this can be applied to BNT162-b2 low dose vaccine regimens to predict protection against COVID-19.

CoPEs allow the concept behind correlates of protection to be applied earlier, as it does not require the extensive research required to define a CoP. This has implications for expeditiously approving treatments against diseases in which the CoP is not yet defined.
